# SYSTEMATIC REVIEW PSYCHOEDUCATION OF ELDERLY WITH DEPRESSION IN PUBLIC HEALTH: PHILOSOPHICAL STUDY

**DOI:** 10.1101/2022.09.27.22280394

**Authors:** Tri Nurhidayati, Moses Glorino Rumambo Pandin, Cindy Nadya

## Abstract

**Introduction:** The increasing number of elderly is a new challenge for the government and health care. The elderly can experience psychological disorders, one of which is depression. The phenomenon of depression that occurs can make sadness and affect the risk of suicide in the elderly. The lack of integration between mental health and primary healthcare services and the shortage of mental health specialists in the public health system contribute to underdiagnosis and undertreatment of depression.One of the strategy to reduce depression is psychoeducation. The purpose of this study was to describe appropriate psychoeducation for elderly people with depression in public health.

**Methodology:** this research design is a literature review, and article using 4 databases, science direct, ProQuest, PubMed, and CINHAL. 16 Article reviewed from 2018 to 2022. The keywords used in the literature search were: 1) psychoeducatioan AND depression AND elderly OR geriatric 2) Depression AND elderly OR geriatric. Protocol and evaluation of literature review using PRISMA checklist and Joanna Briggs Institute Guideline.

**Results:** Psychoeducation can be carried out with various themes, namely providing health information for the elderly such as hypertension and diabetes mellitus, maintaining daily routines, stress management, solving depression problems, social skills, breathing exercises, and information on drug side effects. Psychoeducation is more effective with involving friends, family, and the community.

**Conclusion:** Depression among the elderly will become the world’s problem in the future. Therefore, the quality of elderly depression care needs to be improved. Psychoeducation is more effective by involving family, friends and social environment. Hence, specific psychoeducation for elderly depression can be one of the standards in nursing competencies.

## INTRODUCTION

Aging is a natural process in every human and makes physical and psychosocial changes [1]. The prevalence of depression in the elderly in the world is around 8-15% in women, which is twice as many as in men with a ratio of 14.1: 8.6. In Indonesia, according to the National Commission for the Protection of Older Persons (2010), the prevalence of depression in the elderly is around 30% [1]. Psychosocial disorders that occur in the elderly include depression [2]. Depression in the elderly is characterized by sadness, little interest in activities, and the inability to feel happiness, thus affecting the risk of suicide. The onset of depression in the elderly is often associated with psychological factors, including personality type and interpersonal relationships, including social support. One of the latest therapeutic findings for depressed patients is family psychoeducation which has been proven to be effective and cost-effective for the treatment of patients with major depression in a relatively long period of time [3].

For depression problems experienced by the elderly, the intervention given to the elderly who are depressed is by providing psychoeducation[2]. Psychoeducation is health education aimed at helping people with physical or mental disorders overcome their mental problems. The therapy can be in the form of passive psychoeducation such as providing information that can be given in brochures, emails, or websites and can also be in the form of active psychoeducation in the form of counseling or personal or community health education. Therefore, it is important for individuals and groups as well as families to be able to design care and treatment.[4]

In terms of psychological health, these interventions play an important role as a preventative measure and should be involved with current mental health service delivery[5]. There have been various studies focused on the setting of psychoeducation as an antidote to depression that have previously been carried out [3]. Depression in the elderly requires serious treatment because it can significantly affect health and life.[6] Depression in the elderly will affect the physical activity and personal satisfaction of the elderly. Depression in the elderly can cause physical, mental, emotional, and social changes. [7]. With long goals will generally experience a decrease in personal satisfaction. Thus, researchers are interested in conducting research to address the problem of depression in the elderly. The purpose of this study was to use a literature review approach, namely to understand psychoeducation for depression in the elderly.

## METHODS

The design of this study is a literature review of psychoeducation for elderly people with depression

### Study Protocol

The protocol and evaluation of the literatur review used PRISMA checklist, and the assessment guide used Joanna Briggs Institute Guideline (JBI) to determine the quality of the articles according to the theme

### Article Search Strategy

The data used in this study is based on previous research data. The literature search was carried out from using four databased: Proquest, Pubmed, CINAHL and Science Direct

### Inclusion Criteria

The literature studied were articles published 2018-2022 The article search strategy was developed trough keywords terms in the relevant abstract and title. Keywords in this literature review are adjusted to the Medical Subject Heading (MesSH) which consist psychoeducation, elderly, depression. The keywords used in the literature search were: 1)psychoeducation AND Depression AND elderly OR geriatric 2) Depression And elderly OR geriatric. The strategy used to find articles is using the PICOS framework. Inclusion criteria determined based on articles published in 2018 to 2022, article used in English, access full text, both quantitative and qualitative research.

**Fig 1.**
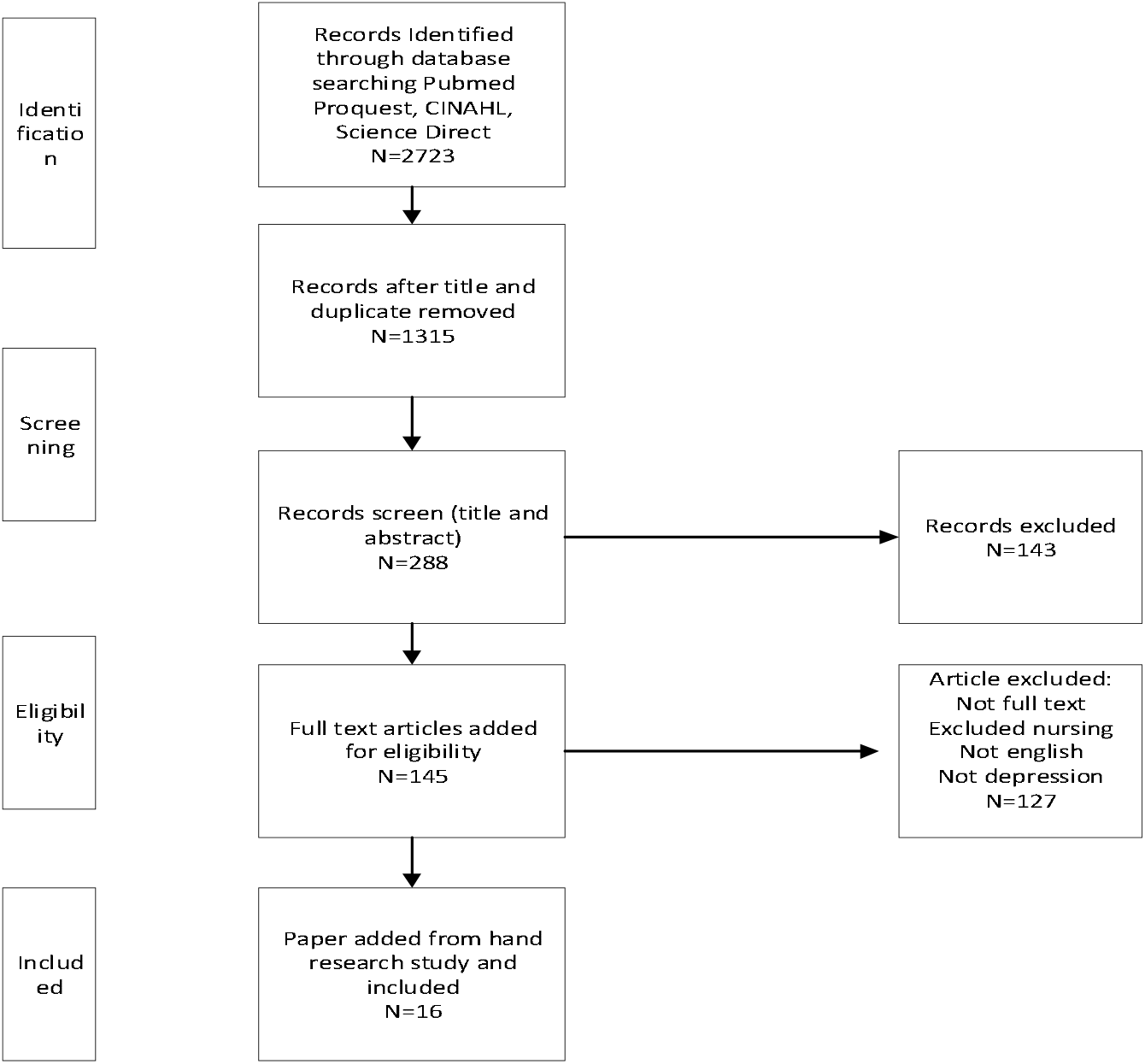
PRISMA Flow Diagram

**Table 1.**
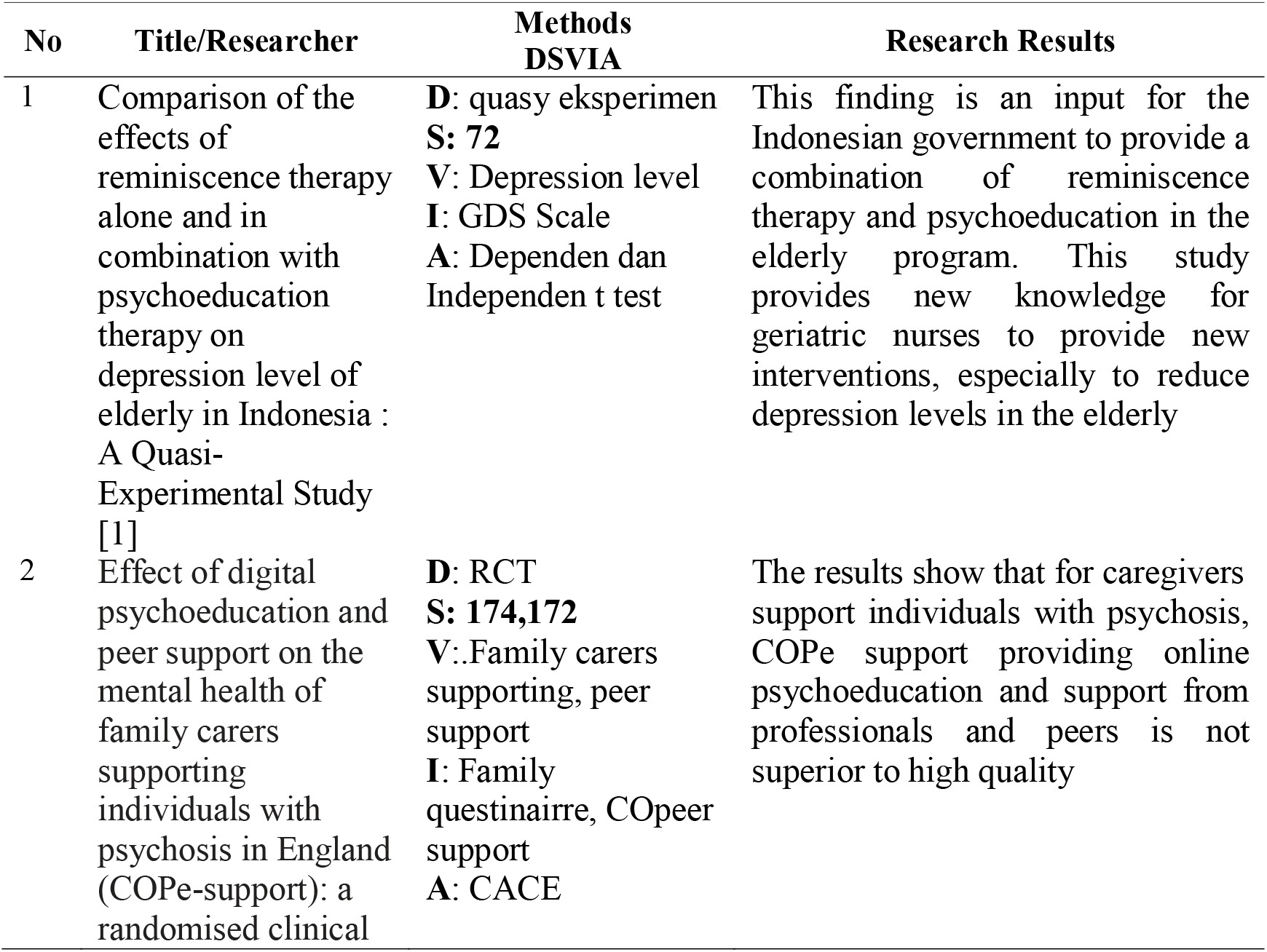
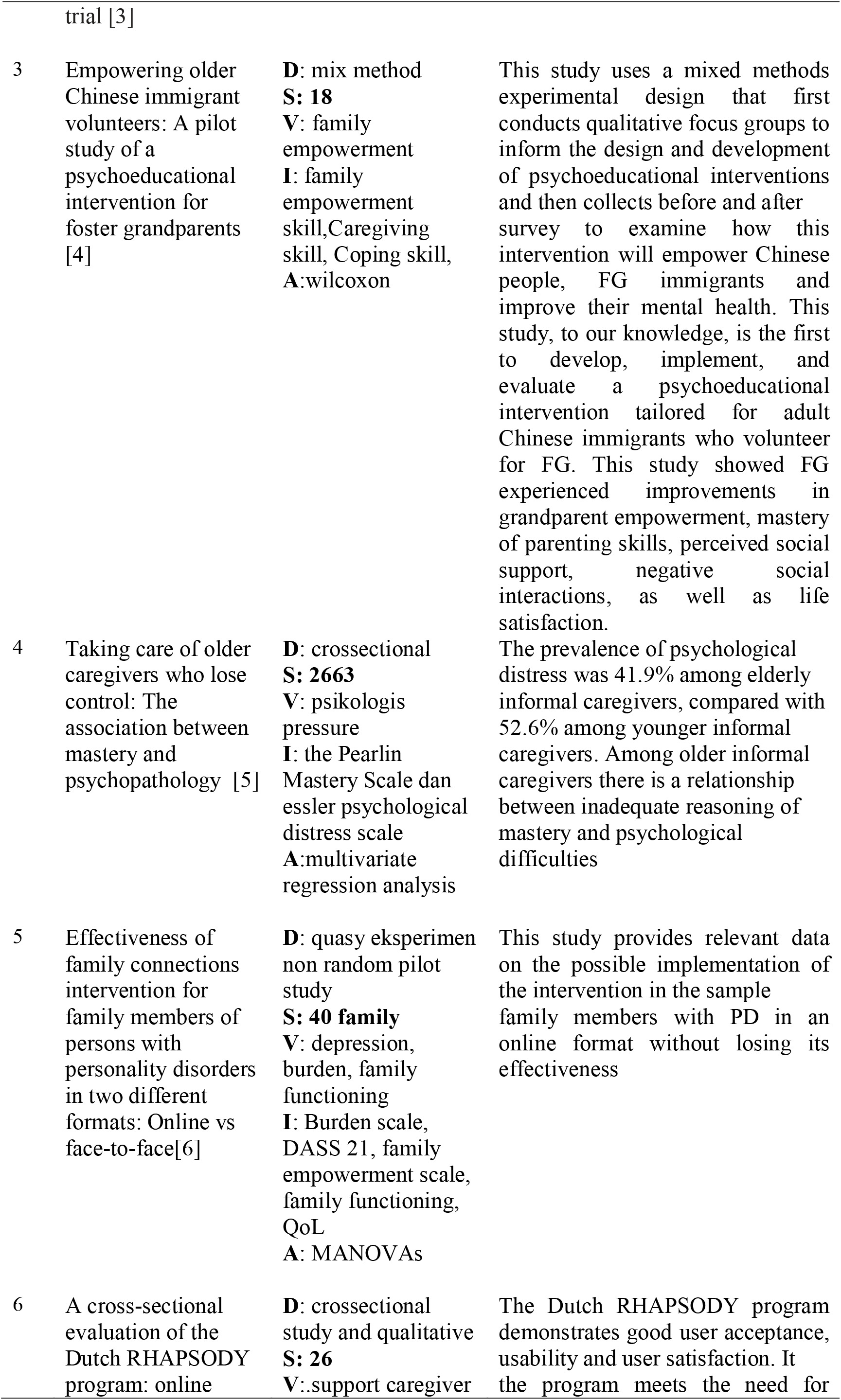
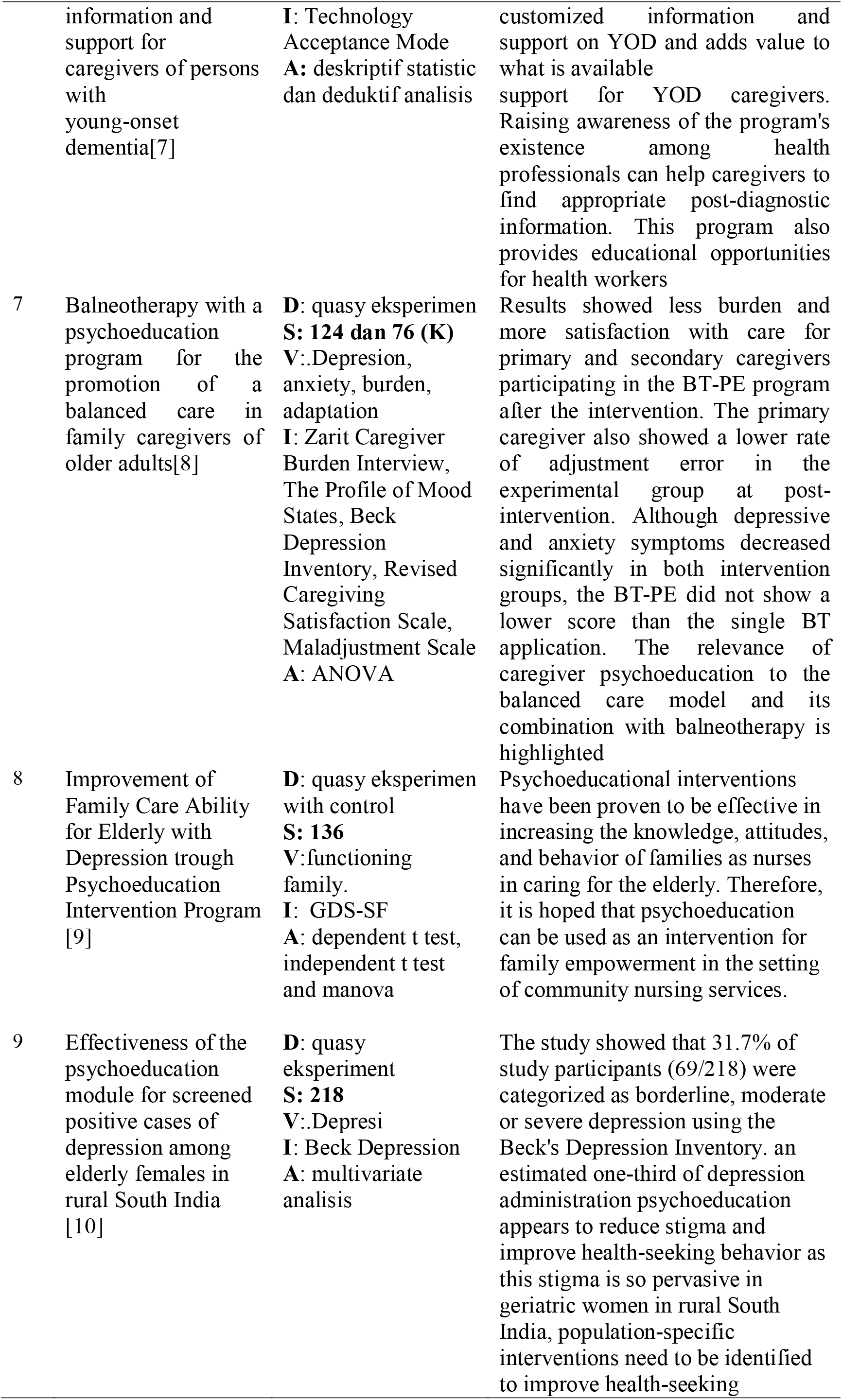
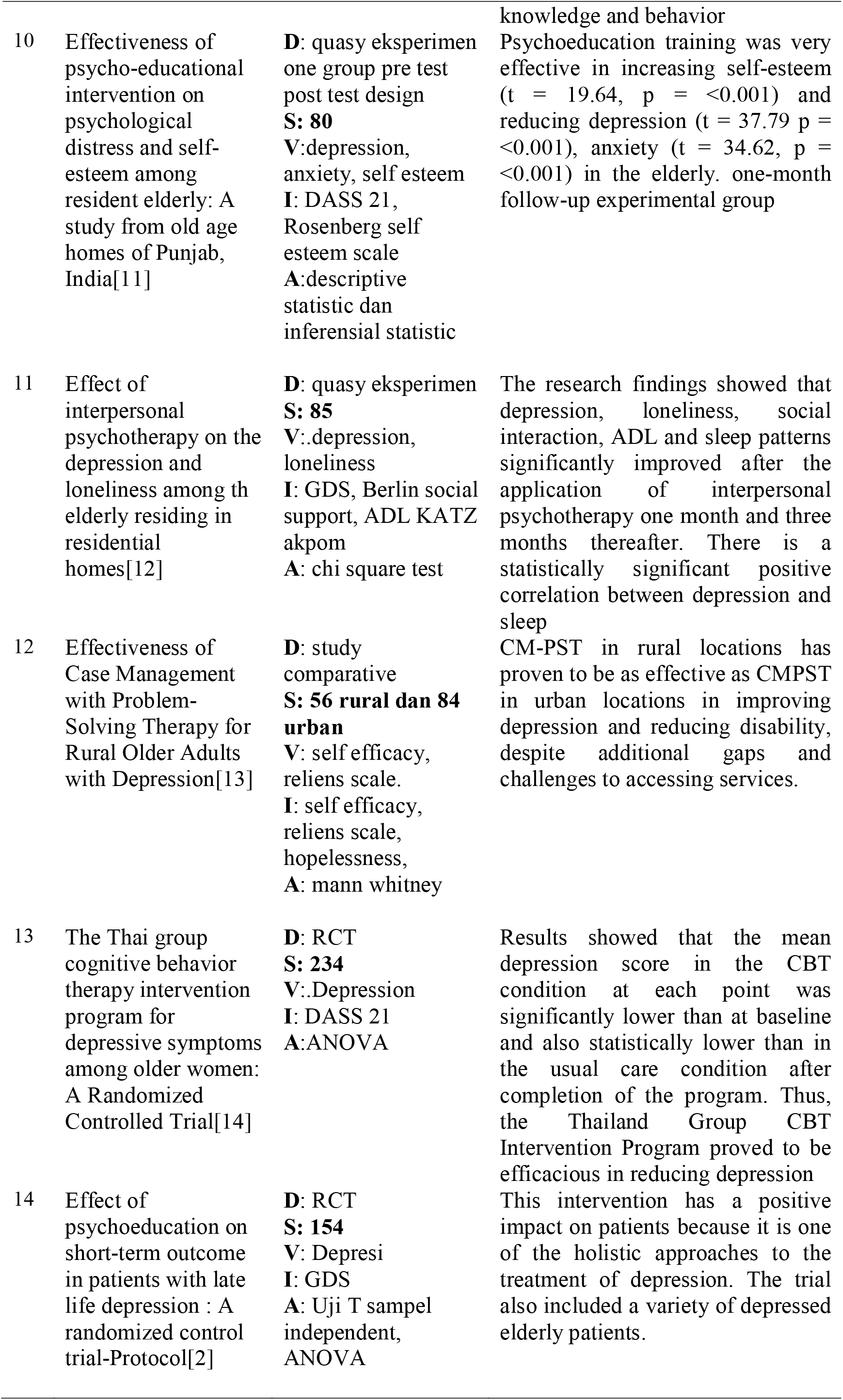
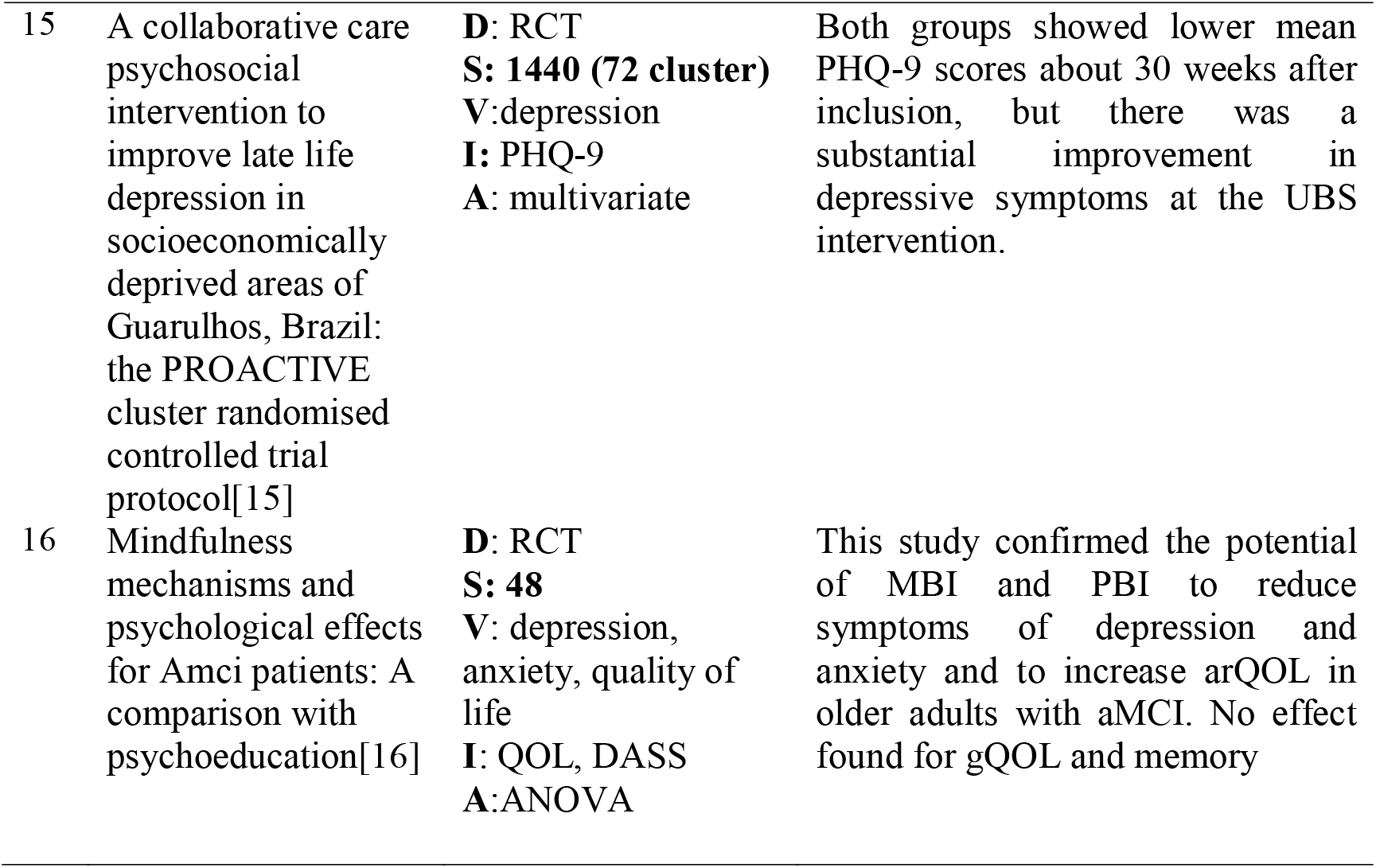
Data Synthesis.

## RESULTS AND DISCUSSION

### 1. Theme of a Depression Psychoeducation Program in the Elderly

This psychoeducational program focuses on implementing interventions that teach the elderly about 1) Health education about: personal hygiene; 2) Information on measures to enhance immunity and management; 3) How to maintain a daily routine; 4) How to maintain health, fun activities, social skills; 5) Health education about Sleep Hygiene; 6) Self-esteem; 7) The role of physical activity and diet in stress management; 8) Diabetes management; 9) Pain management; 10) Breathing exercises; 11) Depression problem solving, behavioral activity, and behavioral cognitive perspective on depression; 12) Management of acidity; 13) Management of Hypertension; 14) Side effects of self-medication. Psychoeducation programs provide the information and skills needed by the elderly to assist them in maintaining physical, mental, social, and emotional health as they age [20]. The psychoeducation intervention program is a form of family empowerment, a strategy to provide information to families about illness and how to treat it by training in coping skills, controlling tension and stress, and interpersonal communication skills. The results show that the psychoeducational intervention program has been shown to be effective in increasing self-esteem, increasing family knowledge, attitudes, and behavior in caring for the elderly, and then reducing the level of depression and loneliness [17][20][15][23]. The following are some of the psychoeducation carried out in the research conducted by the review:

#### a. Interpersonal psychotherapy

Interpersonal Psychotherapy Program (IPT) shows individual interpersonal relationships and current social conditions, for example, problem management to solve problems, encourage and support social support such as building trusting relationships, expressing feelings, increasing self-esteem, and increasing social interaction and interpersonal relationships, increasing independence, and reduce feelings of loneliness and depression. The results of the study showed that the bio-psychosocial condition of the elderly in the form of ADL, sleep patterns, social interactions, feelings of depression, and loneliness of the elderly living in orphanages improved after the application of interpersonal psychotherapy.[1]

#### b. Reminiscence therapy and psychoeducational therapy

Program for giving effect to reminiscence therapy and psychoeducational therapy. Memory therapy that focuses on remembering positive aspects and events that were important to parents in the past, so that the elderly can feel the satisfaction of their lives. While psychoeducational therapy focuses on teaching health education to increase knowledge, and take good care of the elderly. After that, the elderly were given reminiscence therapy and psychoeducational therapy, then a comparison of the two effects of the two therapies was compared. The results showed that reminiscence therapy alone or in combination with psychoeducational therapy had a significant effect on depression rates in older adults.[8]

#### c. Case management with problem solving therapy (CM-PST)

Research on case management with problem solving therapy conducted by (Hollister et al., 2022) in this study focuses on solving a problem. This problem-solving therapy addresses life’s challenges, reduces stress, and facilitates adaptation. This treatment was applied to two groups, the rural elderly group and the urban elderly group. The therapy is given through three stages, the first is psychoeducation, the second is the acquisition of problem solving skills, and the third is the prevention of relapse. The results of this study indicate that CM-PST intervention is an effective intervention for older adults in rural areas, and has been shown to be potentially more effective than CM-PST for older urban adults in improving depression and reducing disability.[18]

#### d. CBT (Cognitive Behavioral Therapy) intervention program

This intervention program consists of: psychoeducation to help understand the causes of depression, case formulation to help describe and explain the problem in a way that is theoretically informed, coherent, meaningful, and leads to effective interventions. The results of the study showed that the CBT intervention program led to a significant reduction in depressive symptoms among older women with mild depression. Participants of the Thailand Group CBT Intervention Program had significantly lower rates of post-intervention depression symptoms than those who received usual care.[19]

### 2. Program Design Equations

Searching articles from 16 journals found programs that can be applied at home to treat the elderly with depression, namely the Psychoeducation Intervention program, Interpersonal Psychotherapy (IPT), case management with problem solving therapy (CM-PST), and the CBT intervention program (Cognitive Behavioral Therapy) from The four programs have something in common, namely involving the family of the elderly, peers, and the social community. In implementing the program, approaches from various aspects that play a role in the success of the program are carried out. This program cannot be effective if one of the parties does not play a role in preventing depression in the elderly and the duration of the intervention is 30 weeks.

### 3. Differences in Program Design

The difference found in the search for 16 journals is the method used. The psychoeducational intervention program focuses on teaching the elderly how to prevent depression with psychoeducational therapy. In providing interpersonal psychotherapy, it shows individual interpersonal relationships and current social conditions, for example, problem management to solve problems, encourage and support social support such as building trusting relationships, expressing feelings, increasing self-esteem, and increasing social interaction and interpersonal relationships, increasing independence, and reduce feelings of loneliness and depression. Meanwhile, case management with problem therapy teaches how to overcome various life challenges, reduce stress, and facilitate life. And for the CBT (Cognitive Behavioral Therapy) intervention program which focuses on cognitive-awareness training programs to help understand the causes of depression, case formulation to help describe and explain the problem in a theoretically informed, coherent, meaningful way, and leads to effective interventions.

## CONCLUSION

Based on research that has been reviewed as many as 16 research journals, it shows that psychoeducation of depression in the elderly has proven to be effective in reducing depression levels in the elderly. Psychoeducation is more effective by involving family, friends and social environment.

## Data Availability

All data produced in the present study are available

## REFERENSI

[1] M. A. El-Bilsha, ‘Effect of Interpersonal Psychotherapy on the Depression and Loneliness among the Elderly Residing in Residential Homes’, Middle East Journal of Age and Ageing, vol. 16, no. 1, pp. 14–25, 2019, doi: 10.5742/mejaa.2019.93620.

[2] L. Lidyana, S. Shelly, and N. Fitria, ‘Pendidikan Kesehatan mengenai Deteksi Dini Depresi dan Penurunan Fungsi Kognitif pada lansia’, Jurnal Abdimas BSI: Jurnal Pengabdian Kepada Masyarakat, vol. 3, no. 1, pp. 12–24, 2020, doi: 10.31294/jabdimas.v3i1.5130.

[3] C. Indriani, Y. S. Hayati, and T. A. Wihastuti, ‘Family Psychoeducation in Reducing the Occurrence of Depression in Elderly: A Systematic Review’, International Journal of Science and Society, vol. 2, no. 3, p. 2020, 2020, [Online]. Available: http://ijsoc.goacademica.com.

[4] D. Bps, ‘Umur Harapan Hidup Saat Lahir (UHH) Tahun 2020-2021’, Badan Pusat Statistik, 2021..

[5] M. R. Alligood, Nursing Theory Utilization & Application, 5th Editio. Elsevier Mosby, 2014.

[6] E. Zheng et al., ‘Health-Related Quality of Life and Its Influencing Factors for Elderly Patients With Hypertension: Evidence From Heilongjiang Province, China’, Frontiers in Public Health, vol. 9, no. March, pp. 1–8, 2021, doi: 10.3389/fpubh.2021.654822.

[7] M. A. Perez-Sousa, P. R. Olivares, J. L. Gonzalez-Guerrero, and N. Gusi, ‘Effects of an exercise program linked to primary care on depression in elderly: fitness as mediator of the improvement’, Quality of Life Research, vol. 29, no. 5, pp. 1239–1246, 2020, doi: 10.1007/s11136-019-02406-3.

[8] Sutinah, ‘COMPARISON of the EFFECTS of REMINISCENCE THERAPY ALONE and in COMBINATION with PSYCHOEDUCATION THERAPY on DEPRESSION LEVEL of ELDERLY in INDONESIA: A QUASI-EXPERIMENTAL STUDY’, Belitung Nursing Journal, vol. 6, no. 1, pp. 1–7, 2020, doi: 10.33546/BNJ.1048.

[9] J. Sin et al., ‘Effect of digital psychoeducation and peer support on the mental health of family carers supporting individuals with psychosis in England (COPe-support): a randomised clinical trial’, The Lancet Digital Health, vol. 4, no. 5, pp. e320–e329, 2022, doi: 10.1016/S2589-7500(22)00031-0.

[10] L. Xu, N. L. Fields, B. C. Tonui, and T. Vasquez-White, ‘Empowering older Chinese immigrant volunteers: A pilot study of a psychoeducational intervention for foster grandparents’, SSM - Mental Health, vol. 2, no. March, p. 100111, 2022, doi: 10.1016/j.ssmmh.2022.100111.

[11] F. M. Kabia, F. el Fakiri, M. Heus, and T. Fassaert, ‘Taking care of older caregivers who lose control: The association between mastery and psychopathology’, Archives of Gerontology and Geriatrics, vol. 101, no. March, p. 104687, 2022, doi: 10.1016/j.archger.2022.104687.

[12] V. Guillén et al., ‘Effectiveness of family connections intervention for family members of persons with personality disorders in two different formats: Online vs face-to-face’, Internet Interventions, vol. 28, no. March, 2022, doi: 10.1016/j.invent.2022.100532.

[13] M. Daemen et al., ‘A cross-sectional evaluation of the Dutch RHAPSODY program: online information and support for caregivers of persons with young-onset dementia’, Internet Interventions, vol. 28, no. March, p. 100530, 2022, doi: 10.1016/j.invent.2022.100530.

[14] C. Noriega, M. D. Ortiz, M. T. Martínez, and J. López, ‘Balneotherapy with a psychoeducation program for the promotion of a balanced care in family caregivers of older adults’, International Journal of Biometeorology, vol. 65, no. 2, pp. 193–203, 2021, doi: 10.1007/s00484-020-02018-4.

[15] N. M. Riasmini, ‘Improvement of Family Care Ability for Elderly with Depression through Psychoeducation Intervention Program’, Jurnal Ilmu dan Teknologi Kesehatan, vol. 8, no. 1, pp. 80–89, 2020, doi: 10.32668/jitek.v8i1.440.

[16] A. Puttur and C. Rao, ‘Effectiveness of the psychoeducation module for screened positive cases of depression among elderly females in rural South India, a Quasi experimental study’, pp. 1–22, 2019.

[17] S. K. Maheshwari, R. Chaturvedi, and P. Sharma, ‘Effectiveness of psychoeducational intervention on psychological distress and self-esteem among resident elderly: A study from old age homes of Punjab, India’, Clinical Epidemiology and Global Health, vol. 11, no. April, p. 100733, 2021, doi: 10.1016/j.cegh.2021.100733.

[18] B. Hollister, R. Crabb, S. Kaplan, M. Brandner, and P. Areán, ‘Effectiveness of Case Management with Problem-Solving Therapy for Rural Older Adults with Depression’, American Journal of Geriatric Psychiatry, vol. 10, no. October, pp. 1083–1092, 2022, doi: 10.1016/j.jagp.2022.03.001.

[19] C. Longchoopol, D. Thapinta, R. Ross, and W. Lertwatthanawilat, ‘The Thai group cognitive behavior therapy intervention program for depressive symptoms among older women: A randomized controlled trial’, Pacific Rim International Journal of Nursing Research, vol. 22, no. 1, pp. 74–85, 2018.

[20] W. Osman Abd El-Fatah, H. Osama Elborie, and O. Ezzat Mahmoud, ‘Psychoeducational and Rehabilitation Intervention guideline for Late-Life Anxiety and Depression among the Older Adults at Geriatric Home’, Egyptian Journal of Health Care, vol. 13, no. 2, pp. 1382–1395, 2022, doi: 10.21608/ejhc.2022.243491.

[21] M. Scazufca et al., ‘Correction to: A collaborative care psychosocial intervention to improve late life depression in socioeconomically deprived areas of Guarulhos, Brazil: the PROACTIVE cluster randomised controlled trial protocol (Trials, (2020), 21, 1, (914), 10.1186/s1306’, Trials, vol. 21, no. 1, pp. 1–14, 2020, doi: 10.1186/s13063-020-04957-0.

[22] E. Larouche, C. Hudon, and S. Goulet, ‘Mindfulness mechanisms and psychological effects for aMCI patients: A comparison with psychoeducation’, Complementary Therapies in Clinical Practice, vol. 34, no. October 2018, pp. 93–104, 2019, doi: 10.1016/j.ctcp.2018.11.008.

[23] A. A. H. Bin Shafie, M. R. B. M. Jailani, N. A. B. A. Miskam, F. A. B. Elias, and H. B. A. Wahab, ‘The Impact of Integrated Psychospiritual Module among the Drug Addicts in Malaysia in Elevating the Psychospiritual and Drug-Related Locus Of Control Level towards the decrease of Relapse Rate’, International Journal of Academic Research in Business and Social Sciences, vol. 8, no. 3, 2018, doi: 10.6007/ijarbss/v8-i3/3929.

